## Supplemental Materials for "The Antidepressant Advisor (ADeSS): A Decision Support System for Antidepressant Treatment for Depression in UK primary care – a feasibility study"

**Short Title: The Antidepressant Advisor**

*[1]Centre for Affective Disorders, Department of Psychological Medicine, King's College London*

*[2]Department of Biostatistics and Health Informatics, King's College London*

*[3]Department of Population Health Sciences, King's College London*

*[4]Department of Health Services & Population Research, King's College London*

*[5] National Service for Affective Disorders, South London and Maudsley NHS Trust*

*Corresponding author:*

*Dr Roland Zahn*
*Centre for Affective Disorders*
*Department of Psychological Medicine*
*Institute of Psychiatry, Psychology & Neuroscience*
*King's College London*
*PO72, 16 De Crespigny Park, London SE5 8AF*
**

### Supplementary Methods

*Intraclass correlation coefficient (ICC) for primary clinical outcome measure*

The intraclass correlation (ICC) describes the extent to which individuals within a cluster are similar to one another. It was calculated by dividing the variance attributable to clustering ($\sigma_u^2$) by the total variance ($\sigma_u^2 + \sigma_e^2$):

$$ICC = \frac{\sigma_u^2}{\sigma_u^2 + \sigma_e^2}$$

In ADeSS, individual patients were clustered at the GP practice level. ICC was calculated for the follow-up measure of the primary clinical outcome (Self-rated Quick Inventory of Depressive Symptomatology sum score; QIDS-SR16) using a mixed effects linear regression model. This is a model that expressed the follow-up score for each individual in terms of an overall intercept ($\beta_0$), a term representing arm differences ($\beta_1$), a term representing the baseline score ($\beta_2$), a cluster-level residual ($u_j$), and an individual-level residual ($\epsilon_{ij}$):

$$\texttt{QIDS\_followup}_{ij} = \beta_0 + \beta_1 \texttt{arm}_{ij} + \beta_2 \texttt{QIDS\_baseline}_{ij} + u_j + \epsilon_{ij}$$

Where $i$ and $j$ refer to patients and GP practices, respectively. Given the small numbers of clusters and patients, confidence intervals for the ICC statistic were calculated with a bootstrap (1000 repetitions, using the bootMer function from the lme4 package in R[1].

*Score for medication side effects during follow-up (FIBSER)*

Side effects were assessed during each week of the treatment period via the mobile app. Patients were invited to complete the FIBSER scale (Frequency, Intensity, Burden of Side Effects Rating[2]) via a notification on their phone.

For five patients who were not using the mobile app, weekly FIBSER scores were additionally collected by the study team via telephone. This was either because the patient did not have a compatible phone or they had technical difficulties using the app.

These data were summarised in two ways. First, we reported data completeness for weekly FIBSER score. Second, we summarised FIBSER (overall and by arm) based on each patient's maximum score for question 3 of the scale ("In the past week, how much have the side effects to your medications for depression interfered with your day-to-day activities?"). which is most relevant for making treatment decisions.

Patients responded on a 7-point scale:

1. No interference with activities
2. Minimal interference with activities
3. Mild interference with activities
4. Moderate interference with activities
5. Marked interference with activities
6. Severe interference with activities
7. Unable to function.

Since some patients responded more often to this questionnaire than others during the treatment period, we summarised this question as each patients' maximum score reported during the 14-week treatment period. This provided a summary that represents each patient's most severe side effects during treatment, without over-representing patients who responded more often. We then calculated the mean (and standard deviation) of the maximum scores across all patients, and by arm.

*Analyses related to economic evaluation*

The aim of the economic component of the study was to ensure that the study procedures needed to complete a future economic evaluation were accurate and feasible.

The cost of the EMIS decision tool was considered as part of the preparatory work for a future economic evaluation. Whilst there were some development costs of the tool, we can consider these sunk costs so not part of any future decision making[3].The decision tool is now part of the national library on EMIS therefore there is no additional cost to subscribers.

Data on service use was collected during the participant interview at baseline, for the six months preceding interview, and at follow-up, for the period since baseline interview, using a modified version of the Adult Service Use Schedule, which had been used in previous studies in depression[4]. In addition, data on referrals to psychiatric services was extracted from EMIS. The EQ5D-3L, used to generate Quality-Adjusted Life Years (QALYs) in a cost-utility analysis was administered in interview at baseline and follow-up[5].

**Supplementary Results**

*Inter-rater reliability on observer-based outcome measures*

Using IBM SPSS25 and a two-way mixed model, with absolute agreement and a single Intraclass Correlation Coefficient (ICC) measure we obtained the following inter-rater reliability estimates:

- MADRS raw scores RZ & PH: ICC = 0.922 based on 16 ratings.
- MADRS raw scores RZ & DF: ICC = 0.809 based on 10 ratings.
- SOFAS raw scores RZ & PH: ICC = 0.630 based on 13 ratings.
- SOFAS raw scores RZ & DF: ICC = 0.962 based on 9 ratings.

*Economic Evaluation*

The utility scores were calculated using standard index values for the United Kingdom[6]. The utility scores and service use data are summarised using descriptive statistics, for all patients and by randomized arm, including mean and standard deviations in Tables A, B and C. The utility scores demonstrate the sensitivity of the measure in this group and the mean scores found here are similar to those for severe (0.39) to moderate depression (0.52) found in larger samples[7]. Service use data collected via self-report using the AD-SUS is detailed for baseline in Table B and follow-up in Table C. At both baseline and follow-up there was no use of secondary care mental health services, with only limited use of secondary care medical health services. The AD-SUS showed that there was substantial use of GP services in the community, but little use of other community-based services other than psychology led services.

### Supplementary Figures

**Figure A|** Daily medication adherence completion via MooDoC mobile app

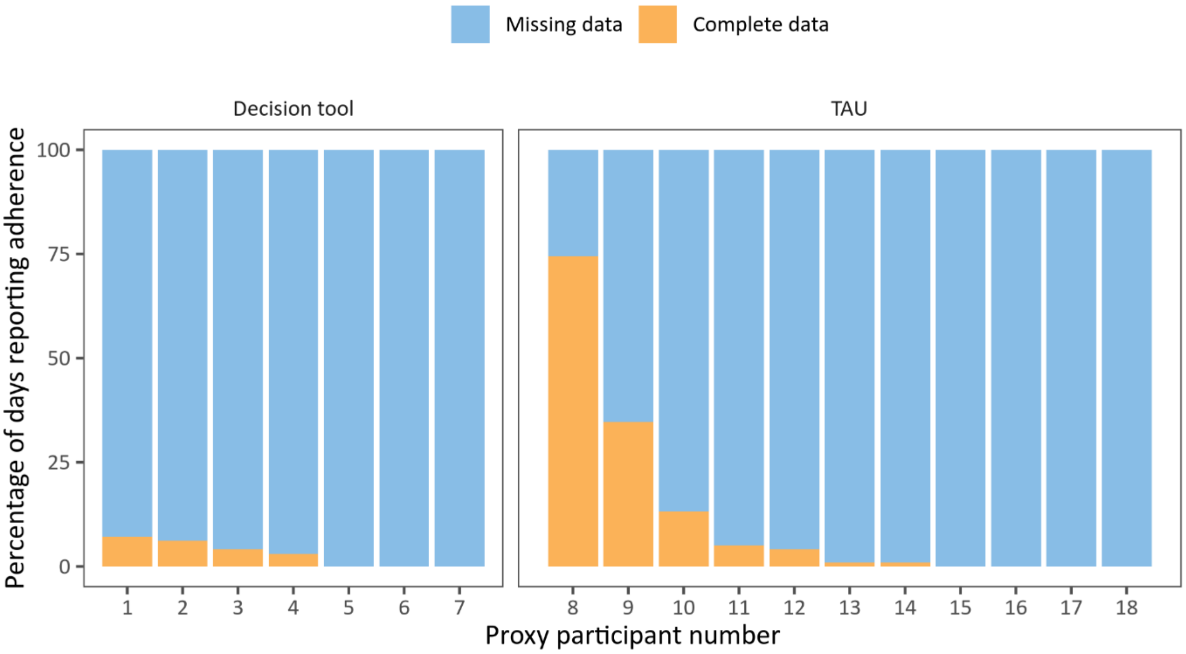


Data completeness of daily medication adherence reports via mobile app (percentage completed vs. percentage missed, during 14-week study period)

### Supplementary Tables

**Table A**| Quality of life

| Interview | Overall | Decision tool | Treatment-as-Usual |
|---|---|---|---|
| Baseline (n=17) | 0.361 (0.362) | 0.439 (0.314) | 0.301 (0.400) |
| Follow-up (n=16) | 0.454 (0.395) | 0.524 (0.384) | 0.400 (0.418) |

Means and standard deviations in parentheses for the EQ-5D-3L[8] Quality of Life measure employed as part of our health economic evaluation.

**Table B**| Self-reported service use at baseline

| | | Overall (n=17) | Decision tool (n=7) | TAU (n=10) |
|---|---|---|---|---|
| Inpatient admission (mental health) | Number (%)with ≥1 admission | 0 (0) | 0 (0) | 0 (0) |
| | Mean (SD) number of nights | 0 (0) | 0 (0) | 0 (0) |
| Inpatient admission (medical) | Number (%) with ≥1 admission | 0 (0) | 0 (0) | 0 (0) |
| | Mean (SD) number of nights | 0 (0) | 0 (0) | 0 (0) |
| Outpatient appointment (mental health) | Number (%) with ≥1 appointment | 0 (0) | 0 (0) | 0 (0) |
| | Mean (SD) number of appointments | 0 (0) | 0 (0) | 0 (0) |
| Outpatient appointment (medical) | Number (%)with ≥1 appointment | 10 (59) | 3 (43) | 7 (70) |
| | Mean (SD) number of appointments | 1.64 (2.06) | 1.43 (2.57) | 1.80 (1.75) |
| Accident and emergency | Number (%) with ≥1 attendance | 4 (24) | 0 (0) | 4 (40) |
| | Mean (SD) number of attendances | 0.24 (0.44) | 0 (0) | 0.40 (0.52) |
| GP – surgery | Number (%) with ≥1 appointment | 14 (72) | 7 (100) | 7 (70) |
| | Mean (SD) number of appointments | 3.41 (4.70) | 5.71 (6.55) | 1.80 (1.93) |
| GP – home | Number (%) with ≥1 visit | 0 (0) | 0 (0) | 0 (0) |

| | | | | |
|---|---|---|---|---|
| | Mean (SD) number of visits | 0 (0) | 0 (0) | 0 (0) |
| GP – telephone | Number (%) with ≥1 appointment | 8 (47) | 2 (29) | 6 (60) |
| | Mean (SD) number of appointments | 1.59 (2.81) | 2.43 (4.24) | 1.00 (1.05) |
| Practice nurse | Number (%) with ≥1 appointment | 6 (35) | 3 (43) | 3 (30) |
| | Mean (SD) number of appointments | 0.82 (1.29) | 0.86 (1.21) | 0.86 (1.21) |
| District nurse | Number (%) with ≥1 appointment | 1 (6) | 0 (0) | 1 (10) |
| | Mean (SD) number of appointments | 0.12 (0.49) | 0 (0) | 0.20 (0.63) |
| Community Psychiatric Nurse | Number (%) with ≥1 appointment | 0 (0) | 0 (0) | 0 (0) |
| | Mean (SD) number of appointments | 0 (0) | 0 (0) | 0 (0) |
| Community psychiatrist | Number (%) with ≥1 appointment | 0 (0) | 0 (0) | 0 (0) |
| | Mean (SD) number of appointments | 0 (0) | 0 (0) | 0 (0) |
| Occupational therapist | Number (%) with ≥1 appointment | 0 (0) | 0 (0) | 0 (0) |
| | Mean (SD) number of appointments | 0 (0) | 0 (0) | 0 (0) |
| Arts therapy | Number (%) with ≥1 appointment | 0 (0) | 0 (0) | 0 (0) |
| | Mean (SD) number of appointments | 0 (0) | 0 (0) | 0 (0) |
| Psychology | Number (%) with ≥1 appointment | 6 (35) | 3 (43) | 3 (30) |
| | Mean (SD) number of appointments | 1.94 (3.38) | 2.00 (2.52) | 1.90 (4.02) |

Service use data collected via self-report using using a modified version of the Adult Service Use Schedule (AD-SUS[9]) at the baseline study visit. TAU=Treatment-as-Usual.

**Table C|** Self-reported service use at follow-up

| | | Overall (n=18) | Decision tool (n=7) | TAU (n=11) |
|---|---|---|---|---|
| Inpatient admission (mental health) | Number (%)with ≥ 1 admission | 0 (0) | 0 (0) | 0 (0) |
| | Mean (SD) number of nights | 0 (0) | 0 (0) | 0 (0) |
| Inpatient admission (medical) | Number (%) with ≥1 admission | 1 (6) | 1 (14) | 0 (0) |
| | Mean (SD) number of nights | 0.06 (0.25) | 0.14 (0.38) | 0 (0) |
| Outpatient appointment (mental health) | Number (%) with ≥1 appointment | 0 (0) | 0 (0) | 0 (0) |
| | Mean (SD) number of appointments | 0 (0) | 0 (0) | 0 (0) |
| Outpatient appointment (medical) | Number (%) with ≥1 appointment | 11 (69) | 5 (71) | 6 (66) |
| | Mean (SD) number of appointments | 1.38 (1.20) | 1.29 (1.11) | 1.44 (1.33) |
| Accident and emergency | Number (%) with ≥1 attendance | 3 (19) | 0 (0) | 3 (33) |
| | Mean (SD) number of attendances | 0.19 (0.40) | 0 (0) | 0.33 (0.50) |
| GP – surgery | Number (%)with ≥1 appointment | 15 (94) | 7 (86) | 8 (89) |
| | Mean (SD) number of appointments | 3.31 (2.98) | 5.14 (3.63) | 1.89 (1.27) |
| GP – home | Number (%) with ≥1 visit | 0 (0) | 0 (0) | 0 (0) |
| | Mean (SD) number of visits | 0 (0) | 0 (0) | 0 (0) |
| GP – telephone | Number(%)with ≥1 appointment | 8 (50) | 4 (57) | 4 (46) |
| | Mean (SD) number of appointments | 1.75 (3.02) | 2.71 (4.27) | 1.00 (1.41) |
| Practice nurse | Number (%)with ≥1 appointment | 7 (44) | 3 (43) | 4 (44) |
| | Mean (SD) number of appointments | 0.63 (0.81) | 0.86 (1.07) | 0.44 (0.53) |
| District nurse | Number (%)with ≥1 appointment | 0 (0) | 0 (0) | 0 (0) |
| | Mean (SD) number of appointments | 0 (0) | 0 (0) | 0 (0) |
| Community Psychiatric Nurse | Number (%)with ≥1 appointment | 0 (0) | 0 (0) | 0 (0) |
| | Mean (SD) number of appointments | 0 (0) | 0 (0) | 0 (0) |

| | | | | |
|---|---|---|---|---|
| Community psychiatrist | Number (%)with ≥1 appointment | 0 (0) | 0 (0) | 0 (0) |
| | Mean (SD) number of appointments | 0 (0) | 0 (0) | 0 (0) |
| Occupational therapist | Number (%)with ≥1 appointment | 2 (6)0.13 | 2 (14) | 0 (0) |
| | Mean (SD) number of appointments | (050) | 0.29 (0.76) | 0 (0) |
| Arts therapy | Number (%)with ≥1 appointment | 0 (0) | 0 (0) | 0 (0) |
| | Mean (SD) number of appointments | 0 (0) | 0 (0) | 0 (0) |
| Psychology | Number (%)with ≥1 appointment | 4 (25) | 3 (43) | 1 (89) |
| | Mean (SD) number of appointments | 1.00 (2.39) | 2.00 (3.42) | 0.22 (0.67) |

Service use data collected via self-report using using a modified version of the Adult Service Use Schedule (AD-SUS[9]) at the four month follow-up study visit. TAU=Treatment-as-Usual.

**Table D|** Number of GP practices enrolled each month during the trial

| | | All GP practices, including those not completing training | | | Enrolled GP practices only, excluding those withdrawn before training | | |
|---|---|---|---|---|---|---|---|
| | | TAU | Decision tool | Total | TAU | Decision tool | Total |
| 2018 | Sep | 1 | 1 | 2 | 1 | 1 | 2 |
| | Oct | 0 | 0 | 0 | 0 | 0 | 0 |
| | Nov | 0 | 0 | 0 | 0 | 0 | 0 |
| | Dec | 1 | 1 | 2 | 1 | 1 | 2 |
| 2019 | Jan | 0 | 0 | 0 | 0 | 0 | 0 |
| | Feb | 2 | 2 | 4 | 2 | 2 | 4 |
| | Mar | 0 | 0 | 0 | 0 | 0 | 0 |
| | Apr | 0 | 0 | 0 | 0 | 0 | 0 |
| | May | 1 | 1 | 2 | 1 | 1 | 2 |
| | Jun | 0 | 0 | 0 | 0 | 0 | 0 |
| | Jul | 0 | 0 | 0 | 0 | 0 | 0 |
| | Aug | 5 | 5 | 10 | 0 | 0 | 0 |
| | Sep | 0 | 0 | 0 | 0 | 0 | 0 |
| | Oct | 0 | 0 | 0 | 0 | 0 | 0 |
| | Nov | 0 | 0 | 0 | 0 | 0 | 0 |
| | Dec | 0 | 0 | 0 | 0 | 0 | 0 |
| 2020 | Jan | 0 | 0 | 0 | 0 | 0 | 0 |
| | Feb | 0 | 0 | 0 | 0 | 0 | 0 |
| | Mar | 0 | 0 | 0 | 0 | 0 | 0 |
| **Total** | | **10** | **10** | **20** | **5** | **5** | **10** |

Number of GP practices recruited and randomised over the course of the trial are summarised. These numbers have been calculated both (i) including all randomised GP practices, including those that were withdrawn shortly after randomisation (left-hand side); and (ii) including only the 10 GP practices that received training and remained in the study (right-hand side).

**Table E|** Responses to GP satisfaction questionnaire from GPs in the 'Decision tool' arm

| Characteristic | N = 5 (%) |
|---|---|
| **How many study patients have you seen?** | |
| 0 | 0 (0%) |
| 1 | 1 (33%) |
| 2-4 | 2 (67%) |
| 5-10 | 0 (0%) |
| >10 | 0 (0%) |
| Missing | 2 |
| **Has the mobile app improved your patients' care, compared to usual practice?** | |
| Very much worse | 0 (0%) |
| Much worse | 0 (0%) |
| Slightly worse | 0 (0%) |
| No change | 0 (0%) |
| Slightly better | 2 (100%) |
| Much better | 0 (0%) |
| Very much better | 0 (0%) |
| Missing | 3 |
| **Has the mobile app affected your workload, compared to your usual practice?** | |
| Very much less work | 0 (0%) |
| Much less work | 0 (0%) |
| Slightly less work | 1 (50%) |
| No change | 0 (0%) |
| Slightly more work | 1 (50%) |
| Much more Work | 0 (0%) |
| Very much more work | 0 (0%) |
| Missing | 3 |
| **How useful did you find the EMIS antidepressant advisor tool?** | |
| Definitely unhelpful | 0 (0%) |
| Probably unhelpful | 0 (0%) |
| Possibly unhelpful | 0 (0%) |
| No opinion | 0 (0%) |
| Possibly helpful | 2 (67%) |
| Probably helpful | 0 (0%) |
| Definitely helpful | 1 (33%) |
| Missing | 2 |
| **Has the EMIS antidepressant advisor tool affected your workload, compared to your usual practice?** | |
| Very much less work | 0 (0%) |
| Much less work | 0 (0%) |
| Slightly less work | 1 (33%) |

| Characteristic | N = 5 (%) |
| --- | --- |
| No change | 1 (33%) |
| Slightly more work | 1 (33%) |
| Much more work | 0 (0%) |
| Very much more work | 0 (0%) |
| Missing | 2 |
| **How easy was it to use the EMIS antidepressant advisor tool?** | |
| Very difficult | 0 (0%) |
| Difficult | 0 (0%) |
| Slightly difficult | 0 (0%) |
| Neutral | 0 (0%) |
| Slightly easy | 1 (33%) |
| Easy | 2 (67%) |
| Very easy | 0 (0%) |
| Missing | 2 |
| **Has the EMIS antidepressant advisor tool improved your patients' care, compared to your usual practice?** | |
| Very much worse | 0 (0%) |
| Much worse | 0 (0%) |
| Slightly worse | 0 (0%) |
| No change | 1 (33%) |
| Slightly better | 2 (67%) |
| Much better | 0 (0%) |
| Very much better | 0 (0%) |
| Missing | 2 |
| **How does this tool compare to other on-screen decision support tools you have used (e.g. Sepsis/QoF alerts)?** | |
| Much worse | 0 (0%) |
| Worse | 0 (0%) |
| Slightly worse | 0 (0%) |
| Equal | 3 (100%) |
| Slightly better | 0 (0%) |
| Better | 0 (0%) |
| Much better | 0 (0%) |
| Missing | 2 |
| **Did you find that you ignored or switched off the EMIS antidepressant advisor tool?** | |
| Always | 0 (0%) |
| Almost always | 0 (0%) |
| Often | 1 (33%) |
| Sometimes | 1 (33%) |
| Almost never | 0 (0%) |
| Never | 1 (33%) |

| Characteristic | N = 5 (%) |
|---|---|
| Missing | 2 |
| **Would you recommend that the EMIS antidepressant advisor tool should be used in future clinical practice?** | |
| Very strongly against | 0 (0%) |
| Strongly against | 0 (0%) |
| Weakly Against | 0 (0%) |
| No opinion | 0 (0%) |
| Weakly support | 3 (100%) |
| Strongly support | 0 (0%) |
| Very strongly support | 0 (0%) |
| Missing | 2 |

Responses to our GP satisfaction questionnaire in GPs using the Antidepressant Advisor Decision Support system implemented into the EMIS electronic health records system.

**Table F|** GP adherence to the algorithm

| GP practice | GP number | Number of patients seen by this GP | None of recommended steps implemented | Less than 50% implemented | More than 50% implemented | Fully implemented |
|---|---|---|---|---|---|---|
| 1 | 1 | 2 | 0 (0) | 0 (0) | 0 (0) | 2 (100) |
| 2 | 1 | 2 | 1 (50) | 0 (0) | 0 (0) | 1 (50) |
| 3 | 1 | 2 | 1 (50) | 0 (0) | 0 (0) | 1 (50) |
| 4 | 1 | 1 | 1 (100) | 0 (0) | 0 (0) | 0 (0) |
| | | Total | 3 | 0 | 0 | 4 |

Adherence to the algorithm as judged by the chief investigator for each GP on the basis of prescription data extracted from EMIS and reported by the patient at follow-up. Reported are number of patients and (percent of all patients seen by GP) in parentheses.

**Table G|** List of adverse event categories

| Arm | Proxy patient ID | Severity | Relationship to study |
| --- | --- | --- | --- |
| Decision tool | 3 | Mild | Possible |
| Decision tool | 7 | Mild | Possible |
| Decision tool | 9 | Mild | Probable |
| Decision tool | 5 | Mild | None |
| TAU | 1 | Mild | Probable |
| TAU | 2 | Mild | None |
| TAU | 4 | Moderate | None |
| TAU | 4 | Moderate | Possible |
| TAU | 8 | Mild | Possible |
| TAU | 6 | Mild | None |

Presented are the adverse events (AEs) recorded during the trial by severity and relationship to the intervention.

**Table H|** Mean scores for FIBSER question 3 during treatment period

| Arm | N avail./N total* | Mean score for FIBSER question 3 (SD) |
| --- | --- | --- |
| Decision tool | 5/7 | 2.2 (1.5) |
| TAU | 8/11 | 2.1 (2.2) |
| Total | 13/18 | 2.2 (1.9) |

*Number of patients for whom a maximum FIBSER score could be derived (i.e. responded at least once during the treatment period), relative to number of patients enrolled. SD=standard deviation. TAU=Treatment-as-usual. FIBSER=Frequency, Intensity, Burden of Side Effects Rating scale[2].

**Table I|** Clinical Global Impression (CGI) scale summarised as a 7-level variable

| | Overall, N = 18[1] | Decision tool, N = 7[1] | TAU, N = 11[1] |
|---|---|---|---|
| Normal | 3 (18%) | 2 (29%) | 1 (10%) |
| Borderline mentally ill | 5 (29%) | 3 (43%) | 2 (20%) |
| Mildly ill | 2 (12%) | 1 (14%) | 1 (10%) |
| Moderately ill | 3 (18%) | 0 (0%) | 3 (30%) |
| Markedly ill | 3 (18%) | 1 (14%) | 2 (20%) |
| Severely ill | 1 (5.9%) | 0 (0%) | 1 (10%) |
| Among the most extremely ill patients | 0 (0%) | 0 (0%) | 0 (0%) |
| Did not attend follow-up | 1 | 0 | 1 |

Clinical Global Impression (CGI; researcher assessed) at follow-up. TAU=Treatment-as-usual. Reported are number of patients and (percent of all patients seen by GP) in parentheses.

**Table J|** Percentage of scheduled GP appointments attended during treatment period

| | Decision tool | TAU | Total |
|---|---|---|---|
| Number of appointments attended, Median [IQR] | 10.5 [6.2, 12.5] | 6.0 [6.0, 12.0] | 8.0 [5.5, 12.5] |
| Number of appointments missed, Median [IQR] | 0.0 [0.0, 0.0] | 0.0 [0.0, 0.0] | 0.0 [0.0, 0.0] |
| Percentage of appointments attended, Mean [SD] | 100.0 [0.0] | 99.5 [1.5] | 99.7 [1.2] |
| Number of patients with EMIS data/ number enrolled* | 6/7 | 9/11 | 15/18 |

*This represents the number of patients for whom data could be extract from EMIS. Of the three patients with missing data, one of these refers to the patient who failed to attend their follow-up interview. TAU=Treatment-as-usual. SD=standard deviation. IQR=Interquartile Range.
