## Supplementary material for "The Antidepressant Advisor (ADeSS): A Decision Support System for Antidepressant Treatment for Depression in UK primary care – a feasibility study": Consort Extension for Feasibility Trials

ADEeSS-CONSORT 2010 checklist of information

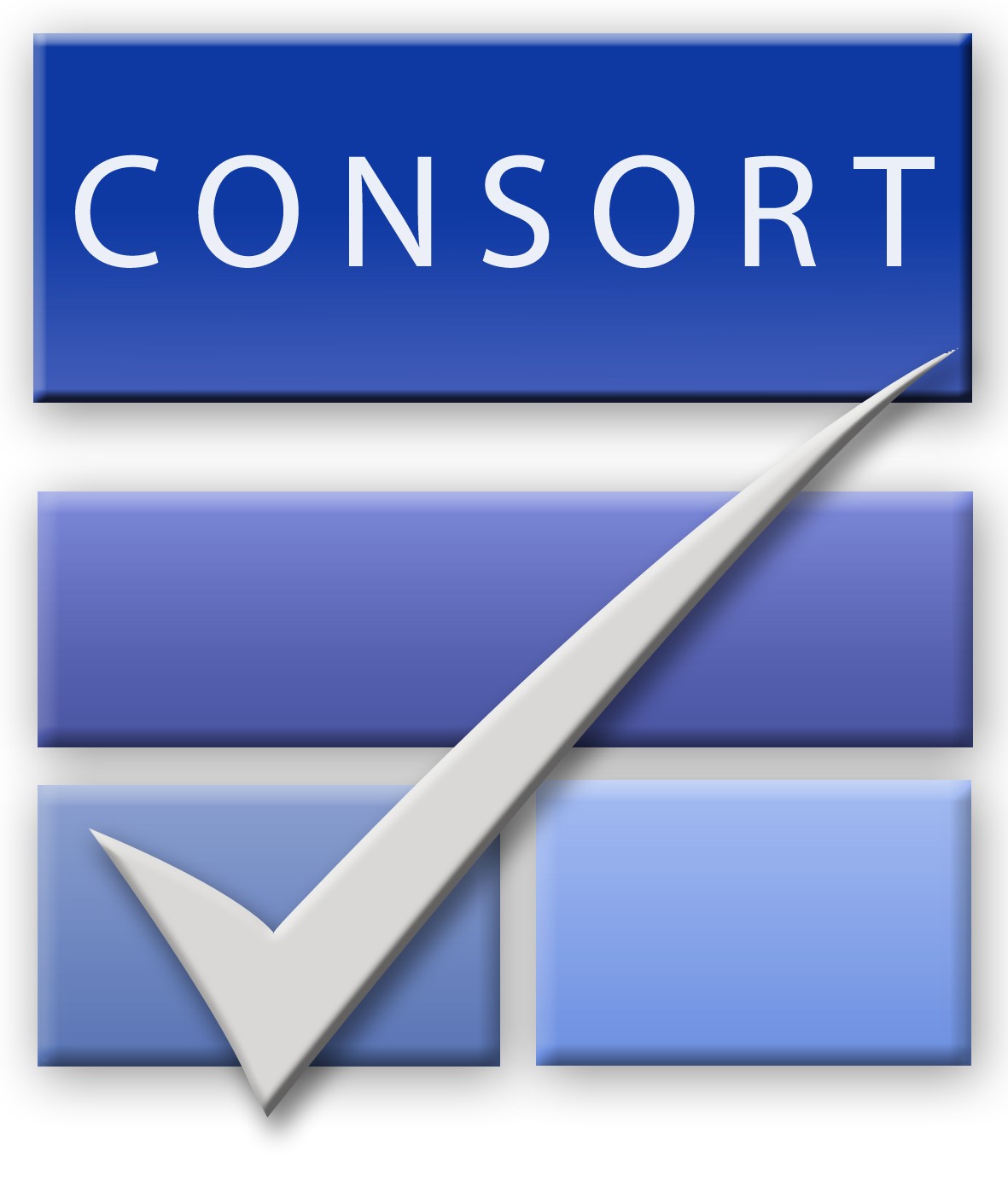

to include when reporting a pilot or feasibility trial*

(also refers to published protocol paper Harrison et al .2020: <http://dx.doi.org/10.1136/bmjopen-2019-035905>)

| Section/Topic | Item No | Checklist item | Reported on page No |
| --- | --- | --- | --- |
| Title and abstract | | | |
|  | 1a | Identification as a pilot or feasibility randomised trial in the title | Title page |
|  | 1b | Structured summary of pilot trial design, methods, results, and conclusions (for specific guidance see CONSORT abstract extension for pilot trials) | Abstract |
| Introduction | | | |
| Background and objectives | 2a | Scientific background and explanation of rationale for future definitive trial, and reasons for randomised pilot trial | P5 |
|  | 2b | Specific objectives or research questions for pilot trial | P6 |
| Methods | | | |
| Trial design | 3a | Description of pilot trial design (such as parallel, factorial) including allocation ratio | P6 |
|  | 3b | Important changes to methods after pilot trial commencement (such as eligibility criteria), with reasons | N/A |
| Participants | 4a | Eligibility criteria for participants | P6-7 |
|  | 4b | Settings and locations where the data were collected | P8 |
|  | 4c | How participants were identified and consented | P8 |
| Interventions | 5 | The interventions for each group with sufficient details to allow replication, including how and when they were actually administered | p10-11 & Harrison et al. 2020: p3-4 |
| Outcomes | 6a | Completely defined prespecified assessments or measurements to address each pilot trial objective specified in 2b, including how and when they were assessed | P8-10 |
|  | 6b | Any changes to pilot trial assessments or measurements after the pilot trial commenced, with reasons | N/A |
|  | 6c | If applicable, prespecified criteria used to judge whether, or how, to proceed with future definitive trial | N/A |
| Sample size | 7a | Rationale for numbers in the pilot trial | Harrison et al. (2020): p5 |
|  | 7b | When applicable, explanation of any interim analyses and stopping guidelines | N/A |
| Randomisation: |  |  |  |
| Sequence  generation | 8a | Method used to generate the random allocation sequence | Harrison et al., 2020:p5 |
|  | 8b | Type of randomisation(s); details of any restriction (such as blocking and block size) | Harrison et al., 2020:p5 |
| Allocation  concealment  mechanism | 9 | Mechanism used to implement the random allocation sequence (such as sequentially numbered containers), describing any steps taken to conceal the sequence until interventions were assigned | Harrison et al., 2020:p5 |
| Implementation | 10 | Who generated the random allocation sequence, who enrolled participants, and who assigned participants to interventions | Harrison et al., 2020:p6 |
| Blinding | 11a | If done, who was blinded after assignment to interventions (for example, participants, care providers, those assessing outcomes) and how | Harrison et al., 2020:p6 |
|  | 11b | If relevant, description of the similarity of interventions | N/A |
| Statistical methods | 12 | Methods used to address each pilot trial objective whether qualitative or quantitative | p11-12 |
| Results | | | |
| Participant flow (a diagram is strongly recommended) | 13a | For each group, the numbers of participants who were approached and/or assessed for eligibility, randomly assigned, received intended treatment, and were assessed for each objective | Figure 1 |
|  | 13b | For each group, losses and exclusions after randomisation, together with reasons | Figure 1 |
| Recruitment | 14a | Dates defining the periods of recruitment and follow-up | P8 & p13 |
|  | 14b | Why the pilot trial ended or was stopped | P8 |
| Baseline data | 15 | A table showing baseline demographic and clinical characteristics for each group | Tables 1&2 |
| Numbers analysed | 16 | For each objective, number of participants (denominator) included in each analysis. If relevant, these numbers  should be by randomised group | All our tables report these by group |
| Outcomes and estimation | 17 | For each objective, results including expressions of uncertainty (such as 95% confidence interval) for any  estimates. If relevant, these results should be by randomised group | All tables reporting quantitative outcomes include these (Tables 3&4 in main manuscript) |
| Ancillary analyses | 18 | Results of any other analyses performed that could be used to inform the future definitive trial | p13 |
| Harms | 19 | All important harms or unintended effects in each group (for specific guidance see CONSORT for harms) | p15 |
|  | 19a | If relevant, other important unintended consequences | N/A |
| Discussion | | | |
| Limitations | 20 | Pilot trial limitations, addressing sources of potential bias and remaining uncertainty about feasibility | P23/24 |
| Generalisability | 21 | Generalisability (applicability) of pilot trial methods and findings to future definitive trial and other studies | P24-25 |
| Interpretation | 22 | Interpretation consistent with pilot trial objectives and findings, balancing potential benefits and harms, and  considering other relevant evidence | P20-25 |
|  | 22a | Implications for progression from pilot to future definitive trial, including any proposed amendments | p20-24 |
| Other information | | |  |
| Registration | 23 | Registration number for pilot trial and name of trial registry | Abstract |
| Protocol | 24 | Where the pilot trial protocol can be accessed, if available | Published: Harrison et al.., 2020 |
| Funding | 25 | Sources of funding and other support (such as supply of drugs), role of funders | Funding statement |
|  | 26 | Ethical approval or approval by research review committee, confirmed with reference number | Abstract |

Citation: Eldridge SM, Chan CL, Campbell MJ, Bond CM, Hopewell S, Thabane L, et al. CONSORT 2010 statement: extension to randomised pilot and feasibility trials. BMJ. 2016;355.

*We strongly recommend reading this statement in conjunction with the CONSORT 2010, extension to randomised pilot and feasibility trials, Explanation and Elaboration for important clarifications on all the items. If relevant, we also recommend reading CONSORT extensions for cluster randomised trials, non-inferiority and equivalence trials, non-pharmacological treatments, herbal interventions, and pragmatic trials. Additional extensions are forthcoming: for those and for up to date references relevant to this checklist, see [www.consort-statement.org](http://www.consort-statement.org).
